# Environmental indicator for effective control of COVID-19 spreading

**DOI:** 10.1101/2020.05.12.20099804

**Authors:** Xinbo Lian, Jianping Huang, Li Zhang, Chuwei Liu, Xiaoyue Liu, Lina Wang

**Affiliations:** Collaborative Innovation Center for Western Ecological Safety, College of Atmospheric Sciences, Lanzhou University, Lanzhou, 730000, China; CAS Center for Excellence in Tibetan Plateau Earth Sciences, Beijing 100101,China; Gansu Province Environmental Monitoring Center, Lanzhou, 730000, China

## Abstract

Recently, a novel coronavirus (COVID-19) has caused viral pneumonia worldwide, spreading to more than 200 countries, posing a major threat to international health. To prevent the spread of COVID-19, in this study, we report that the city lockdown measure was an effective way to reduce the number of new cases, and the nitrogen dioxide (NO_2_) concentration can be adopted as an environmental lockdown indicator. In China, after strict city lockdown, the average NO_2_ concentration decreased 55.7% (95% confidence interval (CI): 51.5-59.6%) and the total number of newly confirmed cases decreased significantly. Our results also indicate that the global airborne NO_2_ concentration steeply decreased over the vast majority of COVID-19-hit areas based on satellite measurements. We found that the total number of newly confirmed cases reached an inflection point about two weeks after the lockdown. The total number of newly confirmed cases can be reduced by about 50% within 30 days of the lockdown. The stricter lockdown will help newly confirmed cases to decline earlier and more rapidly. Italy, Germany and France are good examples. Our results suggest that NO_2_ satellite measurement can help decision makers effectively monitor control regulations to reduce the spread of COVID-19.

## 1. Introduction

Large-scale COVID-19 viral pneumonia through human-to-human transmission poses a severe and acute public health emergency (Huang et al., 2020a; Li et al., 2020). As the epidemic worsened, most countries imposed city lockdown and quarantine measures to reduce transmission to control the epidemic. The Chinese government has gradually implemented a city-wide quarantine of Wuhan and several surrounding cities as of 23 January, flights and trains to and from Wuhan have been suspended, and public transport has been halted (Cyranoski et al., 2020; Wu et al., 2020). The entire northern Italy was quarantined since 9 March 2020, and three days later the government extended it to the whole country (Paterlini, 2020).The Spanish government declared a 15-day national emergency, starting on 15 March (Legido-Quigley et al., 2020). In the United States, on 19 March, California became the first state to order a lockdown (Baccini et al., 2020). In Germany, since 18 March, 16 states have closed, public gatherings of more than two people have been banned and most shops except supermarkets and pharmacies have closed (Dehning et al., 2020). On 23 March, the British government announced a new nationwide restriction allowing residents to only venture outside when absolutely necessary, e.g., to work, buy necessities (Iacobucci, 2020).

The worldwide lockdown, which was imposed to stop the spread of the novel coronavirus, not only caused an economic downturn but also appeared to result in cleaner air in urban areas usually heavily affected by pollution (Lian et al., 2020; Schiermeier, 2020). The most important measure of the lockdown policy was the reduction of traffic and control personnel flow, and traffic pollution is an important factor influencing air quality and public health. Vehicle exhaust and evaporation emissions are the main emission sources of ozone and secondary particle precursors near the ground in cities and regions (Zhou et al., 2019; Yuan et al., 2013), and the spatial variation of nitrogen dioxide (NO_2_), fine particulate matter (PM_2.5_) and black carbon (BC) may also be significant affected by traffic flow density (Clougherty et al., 2013). A study in Los Angeles showed that nitrogen oxides (NO_x_) were identified as a source of pollution for light vehicles, with NO_2_, NO_x_, carbon dioxide (CO_2_), BC, and fine particle number (PN_fine_) identified as diesel exhaust sources (Tessum et al., 2018; Fan et al., 2018). In South Korea, source analysis studies have shown that there is a high correlation between estimated traffic volume and NO_2_ concentration (Kim et al., 2015). NO_2_ levels can be used as a proxy for exposure to traffic-related composite air pollution and to assess the impact of scenarios designed to reduce traffic-related emissions (Brnnum-Hansen et al., 2018; Johansson et al., 2017).

In this report, we study the parameters of environmental indicators for city lockdown. Using the automatic ground detection data and satellite data to analyze the trend of lockdown and the total confirmed new cases in major cities in China, and using satellite data to further study the impact of lockdown on virus transmission in countries mainly severely affected by the epidemic, in order to help policymakers to formulate effective control measures to reduce the spread of COVID-19.

## 2. Data and measurement

The ground observation daily data were provided by the China National Environmental Monitoring Centre (http://www.cnemc.cn/). The data from January 24, 2020, to February 23, 2020, are selected as the representative data after the lockdown in Hubei, and the data from December 24, 2019, to January 23, 2020, are selected as the representative data before the lockdown (Fig. 1). The NO_2_ ground observation data of China is from 1 January 2020 to 1 March 2020. The average concentration of major cities with severe epidemic diseases was selected as the representative of NO_2_ concentration of China, including Wuhan, Nanchang, Guangzhou, Hangzhou, Changsha, Beijing, Shanghai, Hefei and Zhengzhou (Fig. 3). All monitoring instruments of the air quality automatic monitoring system operate automatically 24 h a day. The monitoring items are PM_2.5_, PM_10_, SO_2_, NO_2_, and CO. The automatic monitoring of PM_2.5_ and PM_10_ adopts the micro-oscillating balance method and the β-absorption method, respectively (ambient air quality standards, GB 3095-2012). SO_2_ was determined by the ultraviolet fluorescence method, NO_2_ by the chemiluminescence method, CO by the nondispersion infrared absorption method and gas filter correlation infrared absorption method.

**Fig 1.**
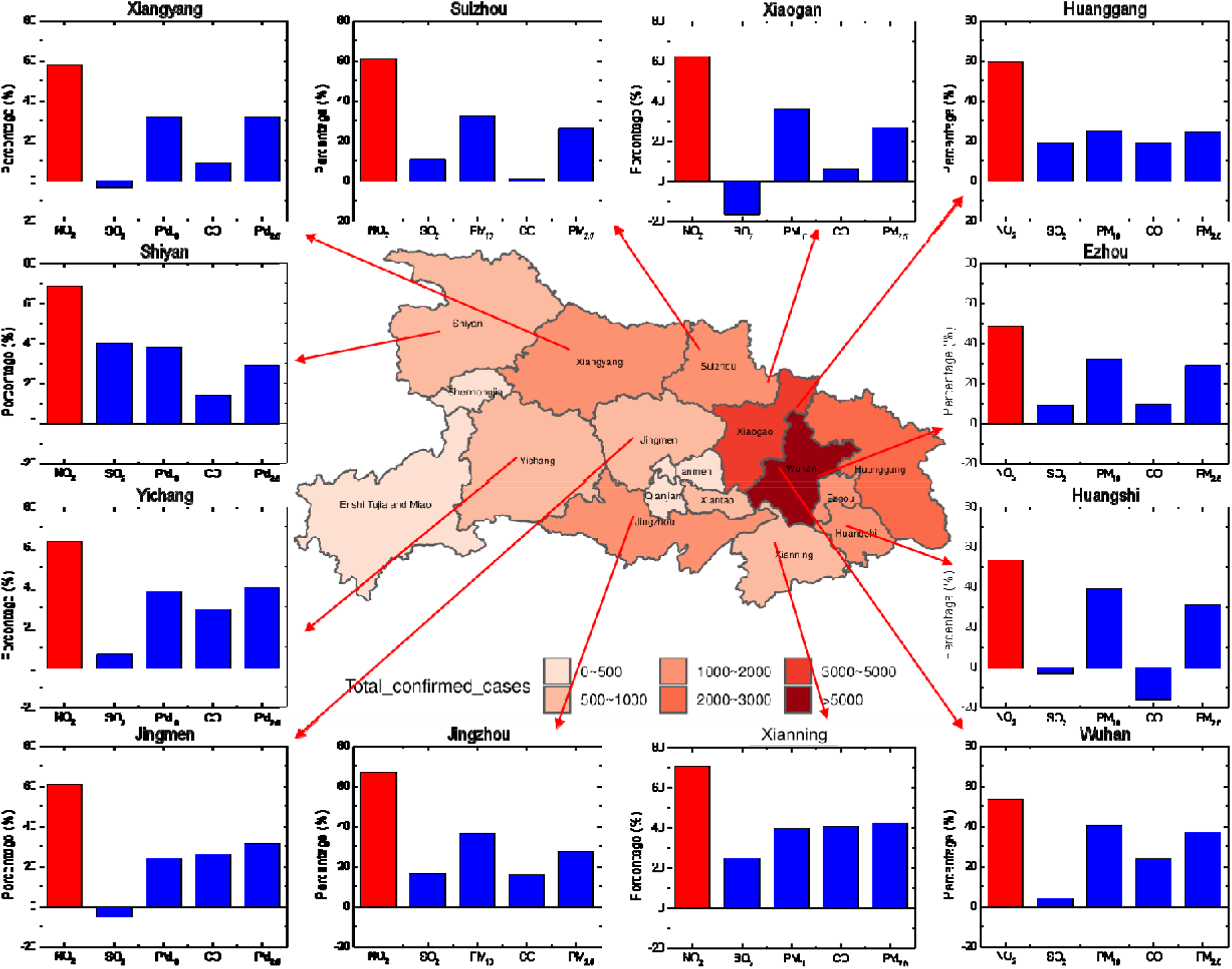
The improvement rate of the major pollutants and the distribution of the accumulated epidemic numbers in each city of Hubei Province after the lockdown. One month after the blockade (23 February 2020), the cumulative number of confirmed cases ranged from 0-46607 (Wuhan), as indicated by the colour scale. The monthly mean improvement rate of NO_2_, SO_2_, PM_10_, CO and PM_2.5_ after the lockdown in the worst virus-affected areas is shown in the histogram, with SO_2_, PM_10_, CO and PM_2.5_ indicated with blue columns, and NO_2_ indicated with red columns. Red columns (NO_2_) are significantly higher than the others, so it can characterize the locking measures.

This paper adopted the level 3 daily global gridded (0.25°×0.25°) nitrogen dioxide product (OMNO2d) provided by the Ozone Monitoring Instrument (OMI) onboard the Aura satellite as the daily NO_2_ data, which can be obtained from GES DISC (https://disc.gsfc.nasa.gov/datasets/OMNO2d_003). The Aura satellite was launched by NASA on July 15, 2004, with its overall objective of monitoring the chemistry and dynamics of the atmosphere from the ground to the mesosphere. The OMI is a nadir-viewing charge-coupled device (CCD) spectrometer onboard the Aura satellite, whose observation band is near-UV/visible. We selected the Column Amount NO_2_ Trop product to calculate the changes in the tropospheric NO_2_ concentration impacted by the control measures in East Asia, Western Europe and North America (Fig. 2):

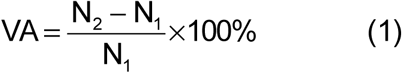

where VA is the relative variation ratio, N1 is the average NO_2_ concentration in the troposphere one month before the lockdown, and N_2_ is the average NO_2_ concentration in the troposphere one month after the lockdown.

**Fig 2.**
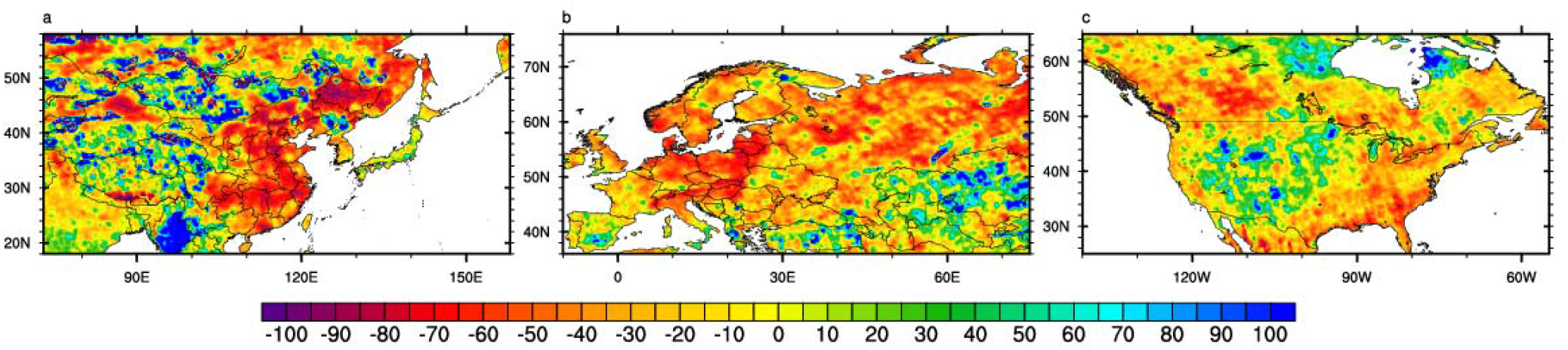
The relative variation in the monthly average tropospheric NO_2_ concentration before and after the lockdown. **a**, Relative variation in East Asia. **b**, Relative variation in Western Europe. **c**, Relative variation in North America. The colour table represents the relative percentage of the monthly average change in NO_2_ after the locksown compared to the before. The red areas indicate that the NO_2_ concentration decreased, and the blue areas indicate that the NO_2_ concentration increased. The lockdown in East Asia, Western Europe and North America began on January 23, March 10, and March 16, respectively. Source: Analysis of data from the NASA Ozone Monitoring Instrument (OMI).

The data from 1 January 2020 to 3 March 2020, were selected to analyze the variation in NO_2_ over time in China (Fig. 3). The NO_2_ satellite data of Italy, Germany, France, the United States, Iran and Switzerland is from the time of the first case in each country to 20 April 2020 (Fig. 4). Due to the satellite orbit, default values occur among the daily data that were determined via piecewise linear interpolation over time. Border data from the US Centers for Disease Control and Prevention (CDC) (https://www.cdc.gov/epiinfo/support/downloads/shapefiles.html) were selected to obtain the borders of each country. To remove the influence of weather factors, a 7-day moving average was calculated. To compare the relative changes among the different countries, the data for each country were standardized.

**Fig 3.**
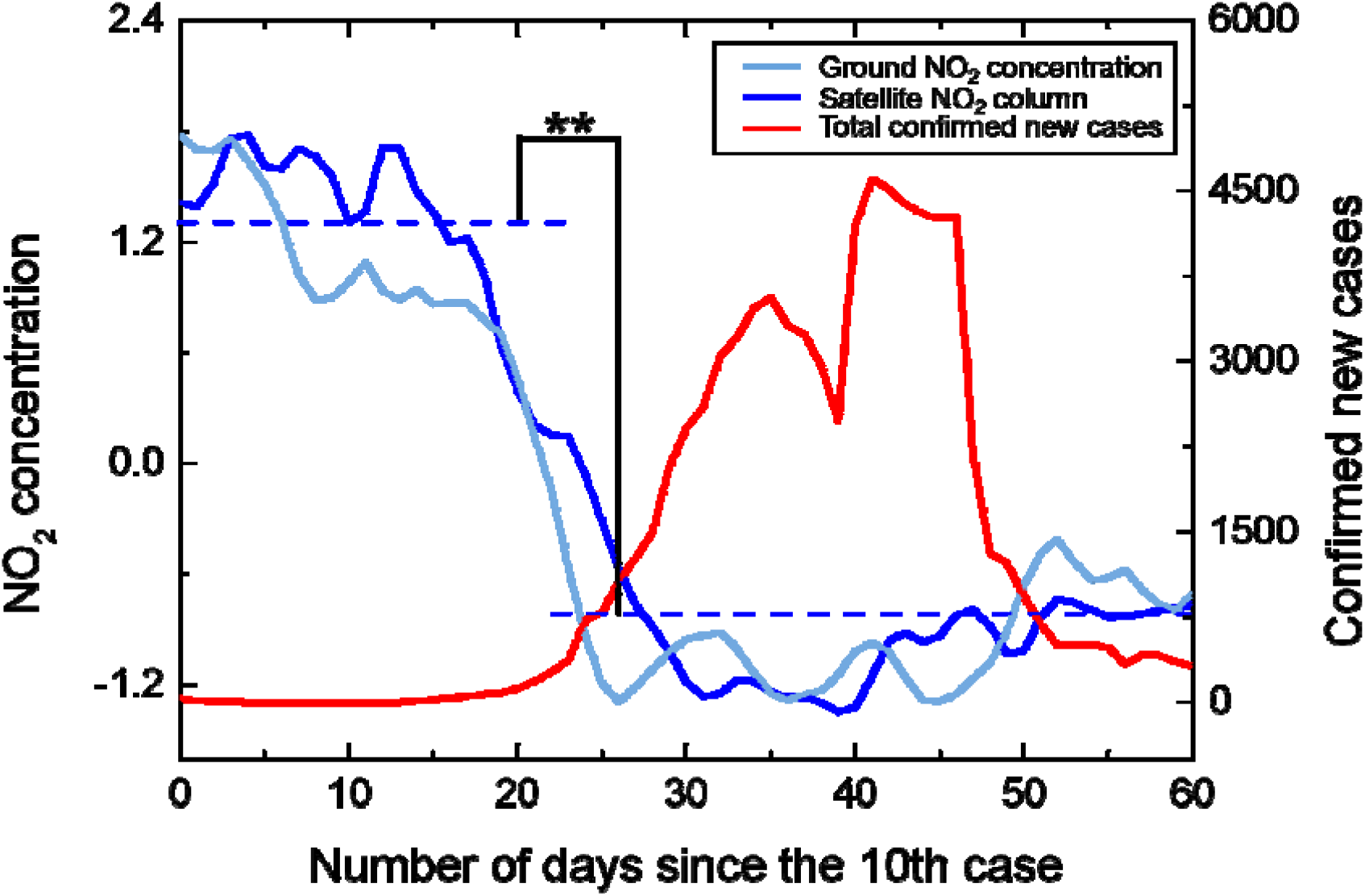
Temporal variation in the NO_2_ concentration and number of new cases in China. The standardized 7-day smooth data of NO_2_ is represented by blue lines, with satellite observations in dark blue and ground observations in light blue. The dotted line indicates the average value of NO_2_ before and after the blockade. The 7-day smooth data of total confirmed new cases is indicated by red lines. ** indicates significant difference at the 0.01 level (bilateral), and * indicates significant difference at the 0.05 level (bilateral). Starting from the lockdown, the NO_2_ concentration dropped significantly, and the number of new cases also showed a downward trend within two weeks.

**Fig 4.**
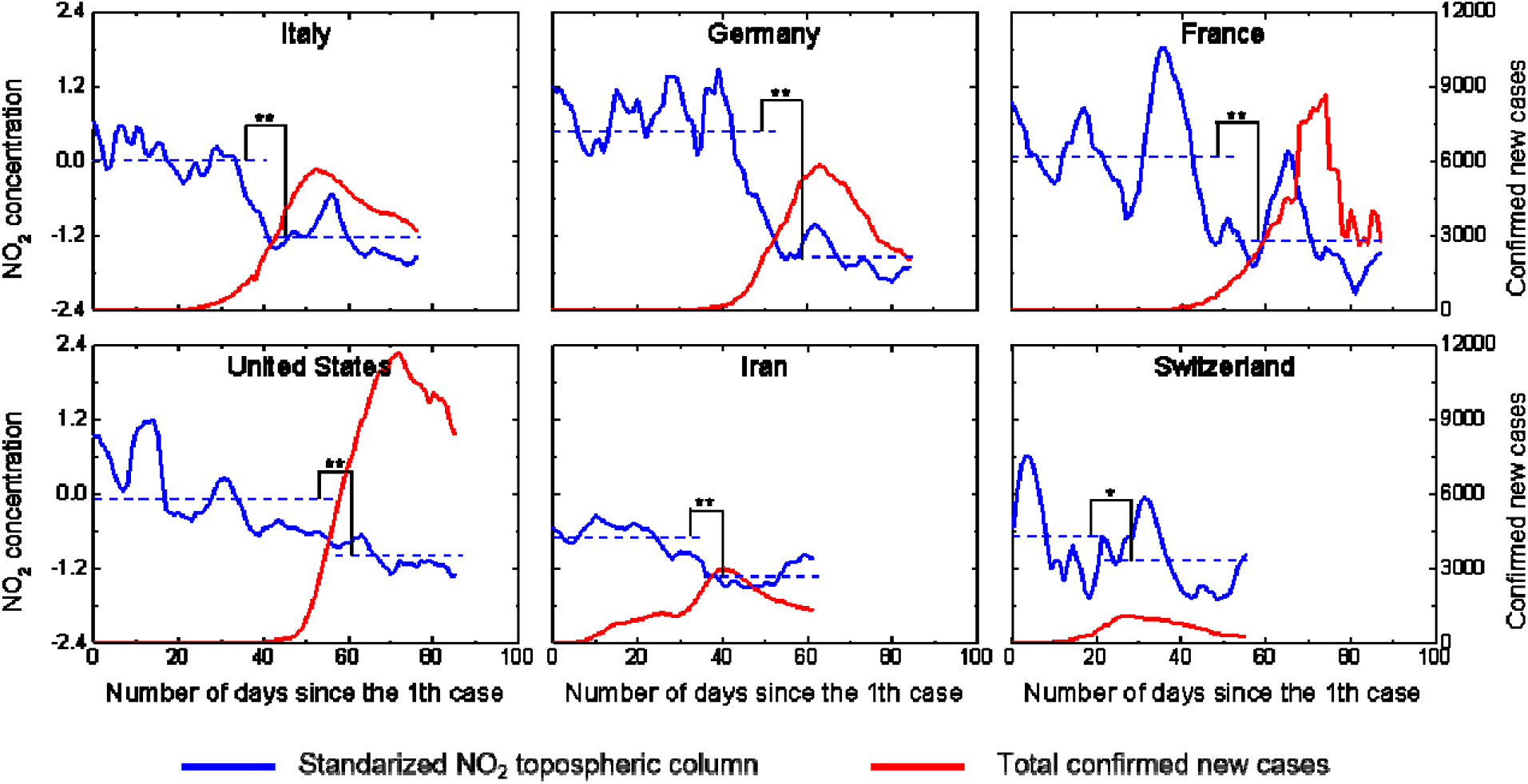
Temporal variation in the NO_2_ concentration and number of new cases in the COVID-19-hit areas. Italy, Germany, France, the United States, Iran and Switzerland were selected as the representatives of COVID-19-hit areas. New York, Washington and California were selected as the representatives of the United States. From the time of the first case in each country to 20 April 2020, the standardized 7-day smooth satellite data of NO_2_ is represented by the blue line. The dotted line indicates the average value of NO_2_ before and after the blockade. The 7-day smooth data of total confirmed new cases is indicated by red line. ** indicates significant difference at the 0.01 level (bilateral), and * indicates significant difference at the 0.05 level (bilateral). The NO_2_ in major countries showed a downward trend. The total confirmed new cases decreased significantly about 15 days after the lockdown.

The daily total number of new confirmed cases in each country and region was obtained from the Center for Systems Science and Engineering (CSSE) of Johns Hopkins University (https://github.com/CSSEGISandData/COVID-19). The daily total number of new confirmed cases in China was retrieved from the Department Earth System Science of the Tsinghua University shared case database (https://cloud.tsinghua.edu.cn/d/335fd08c06204bc49202/).

## 3. Results analysis

### 3.1. A spatial comparison of pollutants before and after lockdown in Hubei Province

Fig. 1 shows the change in pollutant concentration in Hubei Province one month before and after the closure of major cities severely affected by the epidemic. Compared with before the lockdown, NO_2_, sulphur dioxide (SO_2_), particulate matter (PM_10_), carbon monoxide (CO) and PM_2.5_ concentrations all decreased to a certain extent, while NO_2_ experienced the most notable decrease. Since biomass and coal combustion are major SO_2_ and CO sources, they exhibit the lowest rate of improvement (Khalil et al., 1988; Huang et al., 2012). Both the PM_2.5_ and PM_10_ concentrations decreased to a certain extent (31.2% and 34.3%, respectively) as a result of the reduction in fugitive dust, particulate matter and important precursors produced by motor vehicles and factories (Huang et al., 2019). The monthly average PM_2.5_/PM_10_ ratio was 0.81 (95% confidence interval (CI): 0.76-0.86), so PM_2.5_ was the main particle pollutant after lockdown. Exhaust emissions contributed only moderately to local levels of the PM_2.5_ total mass, which were mostly derived from other sources, such as biomass combustion and the remote transmission of secondary particles (Chen et al., 2012). Therefore, the impact of strict traffic control during the lockdown on PM_2.5_ is not notable, and the spatial difference is large, so PM_2.5_ is not suitable as a city lockdown indicator.

Although the NO_2_ emissions per vehicle slightly decreased after the upgrading of the quality standards of petroleum products, the notable growth of vehicle ownership increased the proportion of NO_2_ traffic source emissions, in addition, after the implementation of emission standards for coal-fired power plants, multiple technical improvements greatly controlled the NO_2_ emissions from coal-fired sources, which all enhanced the correlation between NO_2_ and city lockdown effect (Liu et al., 2020). The effect of city closure on NO_2_ was significantly greater than that on the other pollutants, with an average concentration reduction of approximately 60.3% (95% CI: 56.8-64.0%), which can be applied as an environmental indicator of the lockdown effect.

### 3.2. Changes of airborne NO_2_ plummets over COVID-19-hit area after lockdown

In East Asia (Fig. 2a), satellite images show that compared to before the blockade, the total emissions of NO_2_ in eastern China have significantly decreased by approximately 56.6%. In South Korea, the monthly NO_2_ emissions have also been reduced by approximately 18.0 %. The local government has implemented the most expansive testing programme and has isolated people infected with the virus without locking down entire cities, and the sharp decrease in NO_2_ may be linked to the reduction in local emissions and pollutant transport from surrounding areas (Han, 2019; Bauwens et al., 2020). Japan has not imposed widespread lockdown policies, and a 4.8% increase in NO_2_ may be linked to emissions from power generation and industrial processes (Han, 2019). In western Europe (Fig. 2b), the monthly NO_2_ concentrations have decreased sharply in Italy by 47.5%, particularly in the north (82.4%), where the outbreak is the most severe. This could be due to the reduction in road traffic and the decrease in economic activities in the industrial heartland as a result of the widespread lockdown policy (Schiermeier, 2020). Other countries such as Germany, Denmark, and Poland also experienced notable reductions. This is consistent with the results of the European Environment Agency (EEA) (EEA, 2020). However, in certain areas, such as northern and southern Spain, the NO_2_ concentration has risen, possibly because of lax closure measures and increased emissions from coal-fired power plants (Curier et al., 2014). In the United States (Fig. 2c), one month after the lockdown, the overall decline in NO_2_ is relatively small. The worst affected states, such as New York, Washington and California, still contain areas with increased NO_2_ concentrations, and the NO_2_ concentration is increasing significantly in the vast midwestern regions that have not yet been locked down.

### 3.3. The temporal evolution of the NO_2_ concentration and the total number of newly confirmed cases in China

After the strict city lockdown, the NO_2_ concentration in the main virus-affected cities in China decreased significantly (Fig. 3). Consistent with the satellite data, the ground monitoring results showed that compared to the conditions before the closure, the monthly average NO_2_ concentration after the lockdown decreased approximately 55.7% (95% CI: 51.5-59.6%). Since the lockdown, the total number of confirmed new cases reaches an inflection point after approximately two weeks (the incubation period of the virus is 14 days), and compared to the period of 1-15 days after the closure, the total number of confirmed new cases in the 16-30 days after the closure has decreased 73.6% (95% CI: 64.9-81.1%). The most significant improvement was recorded in Hangzhou, where the NO_2_ concentration decreased approximately 68.1%, and the total number of confirmed new cases declined the most. Likewise, Zhengzhou, Changsha, Guangzhou, and Nanchang are good examples. Wuhan, the worst virus-affected area in China, also exhibited a downward trend. The total number of confirmed new cases reached 13,436 on February 12 due to the inclusion of clinically diagnosed cases (Wei et al., 2020), resulting in a new delayed peak in the figure.

The national emergency response has delayed the spread of the epidemic and greatly limited its range. The suspension of intra-city public transport, the closure of entertainment venues and the banning of public gatherings have been linked to a reduction in the incidence of cases. Studies have shown that before emergency response initiation, the basic case reproduction number (R_0_) is 3.15, and after intervention measures were implemented in 95% of all places, the average R_0_ value has dropped to 0.04, the total number of actual cases has decreased 96% (Tian et al., 2020).

### 3.4. The temporal evolution of the NO_2_ concentration and the total number of newly confirmed cases in the COVID-19-hit areas globally

After the countries severely affected by COVID-19 implemented strict lockdown measures, satellite data showed a significant decline in NO_2_ emissions, and the total confirmed new cases decreased after two weeks in most areas (Fig. 4). As a result, the strict lockdown of COVID-19-hit areas other than those in China is also effective and easy to implement to prevent the spread of the virus. The lockdown measures might have already prevented tens of thousands of deaths in Europe (Flaxman et al., 2020). In Italy, where the epidemic is widespread, after the lockdown the NO_2_ emissions significantly decreased by an average of 36.6%, and the total confirmed new cases reached an inflection point 12 days later, thus verifying that the spread of the virus was effectively controlled. Studies have shown that 38,000 deaths have been averted in this country due to the implemented intervention measures (Flaxman et al., 2020). The occurrence time of the inflection point is mainly related to the magnitude of NO_2_ decline. In France, NO_2_ declined less (27.1%), and the time for the total number of confirmed new cases to reach an inflection point was delayed. In Germany, the NO_2_ emissions decreased the most, by 54.7%, and the total number of confirmed new cases reached an inflection point within 8 days after lockdown, which occurred earlier than in other countries. For Iran and Switzerland, due to the relatively low number of confirmed cases, with the decline of NO_2_ after the strict control, confirmed new cases also reached the inflection point earlier. In the the worst-affected states, United States, NO_2_ emissions decreased by an average of 43.1% in New York, Washington and California, and the total confirmed new cases dropped significantly, showing signs of easing.

### 3.5. The results after epidemic control in the COVID-19-hit areas globally

Following the implementation of strict epidemic control measures, the newly confirmed cases dropped by more than 50% within 30 days and NO_2_ declined significantly in most countries (Figure 5a). Especially in most countries in Europe and the Western Pacific region, the number of newly confirmed cases dropped more markedly, with an average drop of more than 60%. The newly confirmed cases in Ireland, Austria, Japan fell by more than 80%. The reduction in the number of newly confirmed cases is closely related to the intensity of interventions, and NO_2_ can quantify the effects and intensity of interventions. Among the top 50 countries with the cumulative number of confirmed cases, the stricter the control measures, the higher the rate of decrease in newly confirmed cases (Figure 5b). If the NO_2_ reduction rate is only about 25%, the control effect of government intervention measures on the epidemic situation is not significant (Figure 5b).

**Fig 5.**
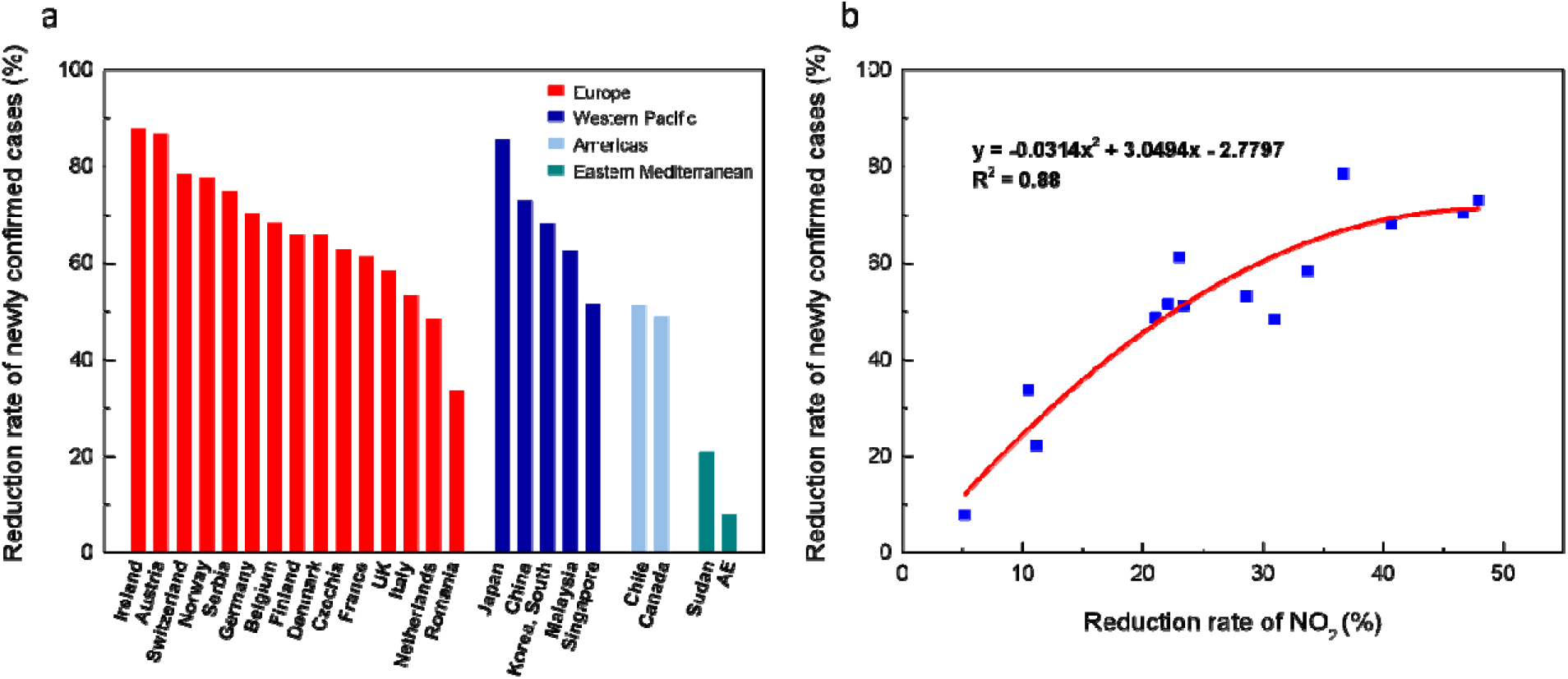
The results of strict epidemic control measures. **a**, The reduction rate of new confirmed cases within 30 days at the NO_2_ mutation point. Red represents Europe, dark blue represents Western Pacific, light blue represents Americas, and green represents Eastern Mediterranean. **b**, Changes in the rate of decline in NO_2_ and the reduction rate of newly confirmed cases after the implementation of strict epidemic control measures.

In Germany, when large public events were canceled, the spreading rate (*λ*) dropped from 0.43 to 0.25. When schools, childcare facilities, and many stores were closed, *λ* dropped to 0.15. Only when a contact ban was imposed by government authorities, *λ* dropped to 0.09, which is lower than the recovery speed, and the exponential growth of new cases turned to decline (Dehning et al., 2020). In Wuhan, China, due to the strict lockdown, the estimated effective reproduction number Rt fell immediately after reaching a peak of 3.82 on January 24 and fell below 1.0 on February 6, 2020 (NO_2_ mutation time) (Pan et al., 2020). In summary, stringent interventions help to reduce newly confirmed cases earlier and more rapidly. NO_2_ can be used as an environmental indicator for quantitative research to assess the efficacy and timing of non-pharmacological interventions to guide current and future responses to global pandemics.

## 4. Discussion

Urbanization and rapid transportation system development accelerate the spread of COVID-19, and only strict containment measures can effectively prevent a second and third outbreak (Huang et al., 2020b; Huang et al, 2020c). The NO_2_ concentration can be considered an inexpensive indicator of virus transmission control. As a result of strict control measures and the rapid implementation of first-level emergency measures, the NO_2_ emissions and total number confirmed new cases significantly decreased in China, especially in the strictly controlled cities. In many European countries, a strict lockdown is also effective and easy to do to prevent the spread of the virus. But there are also areas such as southern and northern Spain and parts of the United States where the NO_2_ level has increased.

Studies have shown that the likelihood of fewer cases in the gradual multi-stage policy is zero, and that such a policy decision implies that the government is willing to risk an increase in COVID19 cases and deaths in exchange for decreased economic and isolation impacts, which may not be desirable from an objective point of view (Karnon, 2020). Although the immediate adoption of a lockdown policy may lead to many people being adversely affected financially, in the short term, the number of new confirmed cases will decline approximately 15 days after policy implementation, and an earlier decline can occur with stricter lockdown measures. International guidance supports a range of mandatory social isolation measures, extensive case detection, and isolation and contact tracing. Compliance with quarantine directives is absolutely critical to saving lives, protecting the most vulnerable in society, and ensuring that the national security system can cope and care for the sick (Iacobucci, 2020). In such cases, an immediate lockdown policy may be preferred, and NO_2_, as an environmental indicator of virus control, can help managers implement effective control measured to curb the spread of COVID-19. In addition, NO_2_ evolution can be used to predict the COVID-19 newly confirmed cases, using the local NO_2_ change rate to reflect the effects of the control measures of governments in various countries. For details, please refer to the Global Prediction System of the COVID-19 Pandemic (http://covid-19.lzu.edu.cn).

## 5. Conclusion

By combining historical COVID-19 data, NO_2_ satellite data and government control measures, we analyzed the quantification and control effects of epidemic intervention measures. The inflection point and reduction rate of newly confirmed cases are closely related to the time and intensity of epidemic control measures. The decline rate of NO_2_ is influenced by the emission characteristics of each country, on the other hand, it is largely related to the intensity and implementation effect of control measures. Therefore, NO_2_ can be used as an environmental indicator to quantify the effectiveness and intensity of interventions and can be incorporated into assessments of the implementation of existing outbreak control measures in countries and into prediction of future scenarios and case numbers. In addition, NO_2_ would help provide policymakers with information on strengthening, easing and selecting appropriate measures to contain COVID-19 and other future global pandemics.

## Data Availability

All data used during the study were provided by a third party. (List items). Direct requests for these materials may be made to the provider.

https://disc.gsfc.nasa.gov/datasets/OMNO2d_003

http://www.cnemc.cn/.

https://github.com/CSSEGISandData/COVID-19

## Acknowledgements

This work was jointly supported by the National Science Foundation of China (41521004) and the Gansu Provincial Special Fund Project for Guiding Scientific and Technological Innovation and Development (Grant No. 2019ZX-06). The authors acknowledge the China National Environmental Monitoring Centre for providing the datasets. http://www.cnemc.cn/.

## Additional information

Competing financial interests: The authors declare no competing financial interests.

## Graphics software

All maps and plots were produced using licensed.

## Notes

### Competing Interest Statement

The authors have declared no competing interest.

### Author Declarations

No patients are involved in this article.This article complies with all relevant ethical guidelines.

## Reference

1. Baccini L, Brodeur A. Explaining governors’ response to the COVID-19 pandemic in the United States. 2020.

2. Bauwens M, Compernolle S, Stavrakou T, Müller JF, Van Gent J, Eskes H, Levelt PF, Vander R, Veefkind JP, Vlietinck J, Yu H, Zehner C. Impact of coronavirus outbreak on NO2 pollution assessed using TROPOMI and OMI observations. Geophys Res Lett 2020; 47.

3. Brnnum-Hansen H, Bender AM, Andersen ZJ, Sørensen J, Bønløkke JH, Boshuizen H, Becker T, Diderichsen F, Loft S. Assessment of impact of traffic-related air pollution on morbidity and mortality in Copenhagen Municipality and the health gain of reduced exposure. Environ Int 2018; 121: 973–980.

4. Chen Y, Liu Q, Geng F, Zhang H, Cai C, Xu T, Ma X, Li H. Vertical distribution of optical and micro-physical properties of ambient aerosols during dry haze periods in Shanghai. Atmos Environ 2012; 50: 50–59.

5. Clougherty JE, Kheirbek I, Eisl HM, Ross Z, Pezeshki G, Gorczynski JE, Johnson S, Markowitz S, Kass D, Matte T. Intra-urban spatial variability in wintertime street-level concentrations of multiple combustion-related air pollutants: The New York City Community Air Survey (NYCCAS). J Expo Sci Env Epid 2013; 23: 232–240.

6. Curier RL, Kranenburg R, Segers AJS, Timmermans RMA, Schaap M. Synergistic use of OMI NO2 tropospheric columns and LOTOS–EUROS to evaluate the NOx emission trends across Europe. Remote Sens of Environ 2014; 149: 58–69.

7. Dehning J, Zierenberg J, Spitzner FP, Wibral M, Neto JP, Wilczek M, Priesemann V. Inferring change points in the spread of COVID-19 reveals the effectiveness of interventions. Science 2020.

8. Fan X, Dawson J, Chen M, Qiu C, Khalizov A. Thermal stability of particle-phase monoethanolamine salts. Environ Sci Tech 2018; 52: 2409–2417.

9. EEA. Air pollution goes down as Europe takes hard measures to combat coronavirus. 2020.

10. Flaxman S, Mishra S, Gandy A. Estimating the number of infections and the impact of nonpharmaceutical interventions on COVID-19 in 11 European countries. Imperial College COVID-19 Response Team 2020.

11. Han KM. Temporal Analysis of OMI-Observed Tropospheric NO2 Columns over East Asia during 2006-2015. Atomos 2019; 10: 658.

12. Huang C, Wang Y, Li X, Ren L, Zhao J, Hu Y, Zhang L, Fan G, Xu J, Gu X, Cheng Z, Yu T, Xia J, Wei Y, Wu W, Xie X, Yin W, Li H, Liu M, Xiao Y, Gao H, Guo L, Xie J, Wang G, Jiang R, Gao Z, Jin Q, Wang J, Cao B. Clinical features of patients infected with 2019 novel coronavirus in Wuhan, China. Lancet (London, England) 2020a.

13. Huang F, Zhou J, Chen N, Li Y, Wu S. Chemical characteristics and source apportionment of PM_2.5_ in Wuhan, China. J Atmos Chem 2019; 76: 245–262.

14. Huang J, Liu X, Zhang Li, Yang K, Chen Y, Huang Z, Liu C, Lian X, Wang D. The amplified second outbreaks of global COVID-19 pandemic. medRxiv 2020b.

15. Huang Q, Cheng S, Perozzi RE. Use of a MM5–CAMx–PSAT Modeling System to Study SO_2_ Source Apportionment in the Beijing Metropolitan Region. Environ Model Assess 2012; 17: 527–538.

16. Huang Z, Huang J, Gu Q, Du P, Liang H, Dong Q. Optimal temperature zone for the dispersal of COVID-19. Sci Total Environ 2020c: 139487.

17. Iacobucci G. Covid-19: UK lockdown is “crucial” to saving lives, say doctors and scientists. Bmj-Brit Med J 2020; 368.

18. Johansson C, Lovenheim B, Schantz P, Wahlgren L, Almstrom P, Markstedt A, Stromgren M, Forsberg B, Sommar J. Impacts on air pollution and health by changing commuting from car to bicycle. Sci Total Environ 2017; 584: 55–63.

19. Karnon J. A Simple Decision Analysis of a Mandatory Lockdown Response to the COVID-19 Pandemic. Appl health econ hea 2020; 18: 329–331.

20. Khalil MAK, Rasmussen, R. A. Carbon Monoxide in an Urban Environment: Application of a Receptor Model for Source Apportionment. Japca 1988; 38: 901–906.

21. Kim Y, Guldmann JM. Land-use regression panel models of NO_2_ concentrations in Seoul, Korea. Atmos Environ 2015; 107: 364–373.

22. Legido-Quigley H, Mateos-Garcia JT, Campos VR, Gea-Sanchez M, Muntaner C, McKee M. The resilience of the Spanish health system against the COVID-19 pandemic. Lancet Public heal 2020; 5: e251–e252.

23. Li Q, Guan X, Wu P, Wang X, Zhou L, Tong Y, Ren R, Kathy SML, Eric HYL, Jessica YW, Xing X, Xiang N. Early Transmission Dynamics in Wuhan, China, of Novel Coronavirus –Infected Pneumonia. New Engl J Med 2020.

24. Lian X, Huang J, Huang R, Liu C, Wang L, Zhang T. Impact of city lockdown on the air quality of COVID-19-hit of Wuhan city. Sci Total Environ 2020: 140556.

25. Liu D, Deng Q, Ren Z, Zhou Z, Song Z, Huang J, Hu R. Variation trends and principal component analysis of nitrogen oxide emissions from motor vehicles in Wuhan City from 2012 to 2017. Sci Total Environ 2020; 704: 134987.

26. Pan A, Liu L, Wang C, Guo H, Hao X, Wang Q, Huang J, He N, Yu H, Lin X. Association of public health interventions with the epidemiology of the COVID-19 outbreak in Wuhan, China. JAMA 2020; 323: 1915–1923.

27. Paterlini M. Lockdown in Italy: personal stories of doing science during the COVID-19 quarantine. Nature 2020.

28. Schiermeier Q. Why pollution is plummeting in some cities - but not others. Nature 2020; 580: 313.

29. Cyranoski D, Silver A. Wuhan scientists: What it’s like to be on lockdown. Nature 2020.

30. Tessum MW, Larson T, Gould TR, Simpson CD, Yost MG, Vedal S. Mobile and Fixed-Site Measurements To Identify Spatial Distributions of Traffic-Related Pollution Sources in Los Angeles. Environ Sci Tech 2018; 52: 2844–2853.

31. Tian H, Liu Y, Li Y, Wu C, Chen B, Kraemer MUG, Li B, Cai J, Xu B, Yang Q. An investigation of transmission control measures during the first 50 days of the COVID-19 epidemic in China. Science 2020; 368: 638–642.

32. Wei Y, Wei L, Jiang Y, Shen S, Zhao Y, Hao Y, Du Z, Tang J, Zhang Z, Jiang Q. Implementation of Clinical Diagnostic Criteria and Universal Symptom Survey Contributed to Lower Magnitude and Faster Resolution of the COVID-19 Epidemic in Wuhan. Engineering 2020.

33. Wu JT, Leung K, Leung GM. Nowcasting and forecasting the potential domestic and international spread of the 2019-nCoV outbreak originating in Wuhan, China: a modelling study. Lancet (London, England) 2020.

34. Yuan B, Hu W, Shao M, Wang M, Chen W, Lu S, Zeng L, Hu M. VOC emissions, evolutions and contributions to SOA formation at a receptor site in eastern China. Atmos Chem Phys 2013; 13: 8815–8832.

35. Zhou Z, Tan Q, Liu H, Deng Y, Wu K, Liu C, Zhou X. Emission characteristics and high-resolution spatial and temporal distribution of pollutants from motor vehicles in Chengdu, China. Atmos Pollut Res 2019; 10: 749–75.

